# Curating and translating the evidence about SARS-CoV-2 and COVID-19 for frontline public health and clinical care: The Novel Coronavirus Research Compendium (NCRC)

**DOI:** 10.1101/2021.04.26.21255437

**Authors:** Andrew D. Redd, Lauren Peetluk, Brooke Jarrett, Colleen Hanrahan, Sheree Schwartz, Amrita Rao, Andrew Jaffe, Carli Jones, Chelsea Lutz, Clif McKee, Eshan Patel, Greg Rosen, Henri Garrison Desany, Heather McKay, John Muschelli, Kayte Andersen, Malen A. Link, Nikolas Wada, Prativa Baral, Ruth Young, Denali Boon, M. Kate Grabowski, Emily S. Gurley, the NCRC Team

## Abstract

The public health crisis created by the SARS-CoV-2 pandemic has spurred a deluge of scientific research aimed at informing public health and medical response to the COVID-19 pandemic. However, those working in frontline public health and clinical care had insufficient time to parse the rapidly evolving evidence and use it for decision making. Academics in public health and medicine were well-placed to translate the evidence for use by frontline clinicians and public health practitioners. The Novel Coronavirus Research Compendium (NCRC), a group of >50 faculty and trainees, began in March 2020 with the goal to quickly triage and review the large volume of preprints and peer-reviewed publications on SARS-CoV-2 and COVID-19, and to summarize the most important, novel evidence to inform pandemic response. From April 6, 2020 through January 1, 2021, 54,192 papers and preprints were screened by NCRC teams and 527 were selected for review and uploaded to the NCRC website for public consumption. The majority of papers reviewed were peer-reviewed publications (n=395, 75%), published in 102 journals; 25% (n=132) of papers reviewed were of preprints. The NCRC is a successful model of how academics can support practitioners by translating scientific knowledge into action and help to build capacity among students for this work. This approach could be used for health problems beyond COVID-19, but the effort is resource intensive and may not be sustainable over the long term.

## Introduction

The public health crisis created by the SARS-CoV-2 pandemic has spurred an unprecedented response from the public health and scientific community to generate evidence about transmission, clinical presentation, pathogenesis, and best practices for prevention and mitigation. Between January 30 and April 23, 2020, there were an average of 367 articles published weekly about SARS-CoV-2 with a median submission-to-publication time of six days (Palayew et al. 2020). In comparison, when the World Health Organization declared Ebola as an international emergency in 2019, there were four articles published weekly with a median submission-to-publication time of 15 days. More than 100,000 papers were published on SARS-CoV-2 between December 2019 and December 2020 (https://coronacentral.ai/), more than all papers ever published for infectious diseases such as measles (∼53,000) or Lyme disease (∼24,000)^1^. The pandemic emergency has necessitated such urgent sharing of scientific evidence that many authors have increasingly turned to preprints to share results quickly despite the acceleration in publication speed (Horbach 2020).

The large volume of new evidence about SARS-CoV-2 was produced with the aim of improving our collective medical and public health response; however, this aspiration can only be realized if, at a minimum, the best evidence is seen by practitioners and policy makers at the right time. Early in the pandemic, two barriers to optimal use of emerging evidence became clear. First, clinicians and public health practitioners on the frontlines of the pandemic response did not have the time to keep up with the rapid pace of new knowledge generation. Their time and effort were completely consumed with patient care and ramping up public health responses. Second, given the diverse research disciplines represented in the emerging literature, few individuals are likely to have the required technical expertise to appropriately evaluate the evidence across broad topic areas and to determine its relevance to clinical or public health practice. It can be even more difficult to assess the strengths and weaknesses of evidence presented in preprints because they, by definition, are often less developed than published papers.

Academics in public health and medicine are well-placed to help facilitate the use of this evidence by frontline clinicians and public health practitioners. They routinely review and critique the scientific literature, and collectively have the technical training to assess strengths and weaknesses of studies across a wide variety of scientific fields.

### Purpose

The Novel Coronavirus Research Compendium (NCRC) began in March 2020, with the goal to quickly triage and review the large volume of preprints and peer-reviewed publications on SARS-CoV-2 and COVID-19, and to summarize the most important, novel evidence to inform health departments, clinicians, and policymakers responsible for pandemic response. Here, we present our process and experiences with this initiative to provide a case study in knowledge translation for pandemic response and to serve as a reference for others considering similar evidence curation efforts for other diseases.

## Methods

### Composition and expertise of teams

The NCRC comprises eight teams focused on clinical presentation of COVID-19, diagnostics, ecology and spillover, epidemiology, disease modeling, non-pharmaceutical interventions, pharmaceutical interventions, and vaccines. Each topic is led by faculty with expertise in that area, supported by other faculty, doctoral trainees, postdoctoral fellows, or medical students. The NCRC is led by a team at Johns Hopkins University and includes >60 individuals from a variety of institutions.

### Identifying and selecting papers for review

The first papers reviewed by the NCRC (n=54) were selected by the NCRC team based on their knowledge of the most important papers published and preprints available through March 30, 2020. From that date forward, manuscript searches were automated. An informationist coordinated with each team to develop eight search queries – one per research area – to identify articles on COVID-19 and SARS-CoV-2. For PubMed, queries are invoked with the *easyPubMed* R package [https://cran.r-project.org/web/packages/easyPubMed/index.html] and reformatted from XML using the *rvest* package (https://cran.r-project.org/web/packages/rvest/index.html) to extract relevant information. For preprints from bioRxiv and medRxiv, papers are obtained from their “COVID-19 SARS-CoV-2 preprints” curated collection (https://connect.biorxiv.org/relate/content/181) using their application programming interface (API) in the JSON format using the *jsonlite* R package (https://cran.r-project.org/web/packages/rvest/index.html). Preprints from SSRN are obtained as daily XML files through the Elsevier Developers API and are processed as above. All preprints are assigned to each of the research areas using predefined search queries. Articles that match multiple research areas are assigned to the group with the fewest papers.

Metadata about papers returned by this process are appended to a Google Sheets spreadsheet that serves as the back-end database for an R Shiny web application, which was developed to be the primary interface for NCRC teams to access and select the papers generated through the automated search results.

In the “triage” process, NCRC members selected papers for in-depth review that (1) contain original research (can include systematic reviews) representing important contributions to our understanding of the pandemic that would be relevant to a practice-based public health audience and/or (2) are widely circulated in the public sphere. Studies are also sometimes identified by members through news reports or social media before the automated process; these are processed outside of the R Shiny application.

### Structure of the reviews and editing

NCRC members read articles selected for review and identified the study population and design, highlighted the major findings, study strengths and limitations, and summarized the added value of the evidence considering what was already known. Each review also included a section called ‘Our Take,’ which provides a capsule evaluation in ∼150 words. Reviews are typically drafted by doctoral trainees and reviewed by faculty before being entered into the R Shiny application. These submitted reviews undergo a second round of faculty editing for clarity and consistency of communication. Final reviews are then automatically posted to the NCRC website (https://ncrc.jhsph.edu/) directly from a Google Sheet, which automatically populates sections. Through a formal collaboration with bioRxiv and medRxiv, NCRC reviews of most preprint papers are also automatically posted onto the preprint’s bioRxiv or medRxiv page.

### Communicating reviews

The NCRC website launched on April 27, 2020. Paper reviews were organized across the eight research areas (with the ability to cross-post to all relevant areas). A search function allows viewers to search for reviews based on words appearing anywhere in the review. On June 15, 2020, each review began to include the date it was published to the NCRC website. Two reviews were of papers eventually retracted due to lack of data reproducibility. For these papers, the phrase “This article has been retracted due to concerns over data veracity” replaced our reviews on the website.

On July 2, 2020, an email newsletter was launched where subscribers received a weekly digest of all new reviews posted to the website. The team has also used Twitter (@JHSPH_NCRC) to highlight noteworthy articles and to post topical threads with an evidence summary and links to multiple related reviews. Popular hashtags (e.g., #COVID19, #SARSCoV2, and #coronavirus) are used to help people who are not yet following @JHSPH_NCRC to find these posts.

### Outcomes

From April 6, 2020 through January 1, 2021, 54,192 papers and preprints were uploaded into the R Shiny application for teams to triage and review. The number of articles uploaded to Shiny increased through August and decreased in the following months (Table 1). The majority of the papers uploaded to Shiny were publications in peer-reviewed journals (76%), while the other 24% were from preprint archives (bioRxiv, medRxiv, SSRN, and Research Square) (Table 1).

**Table 1.**
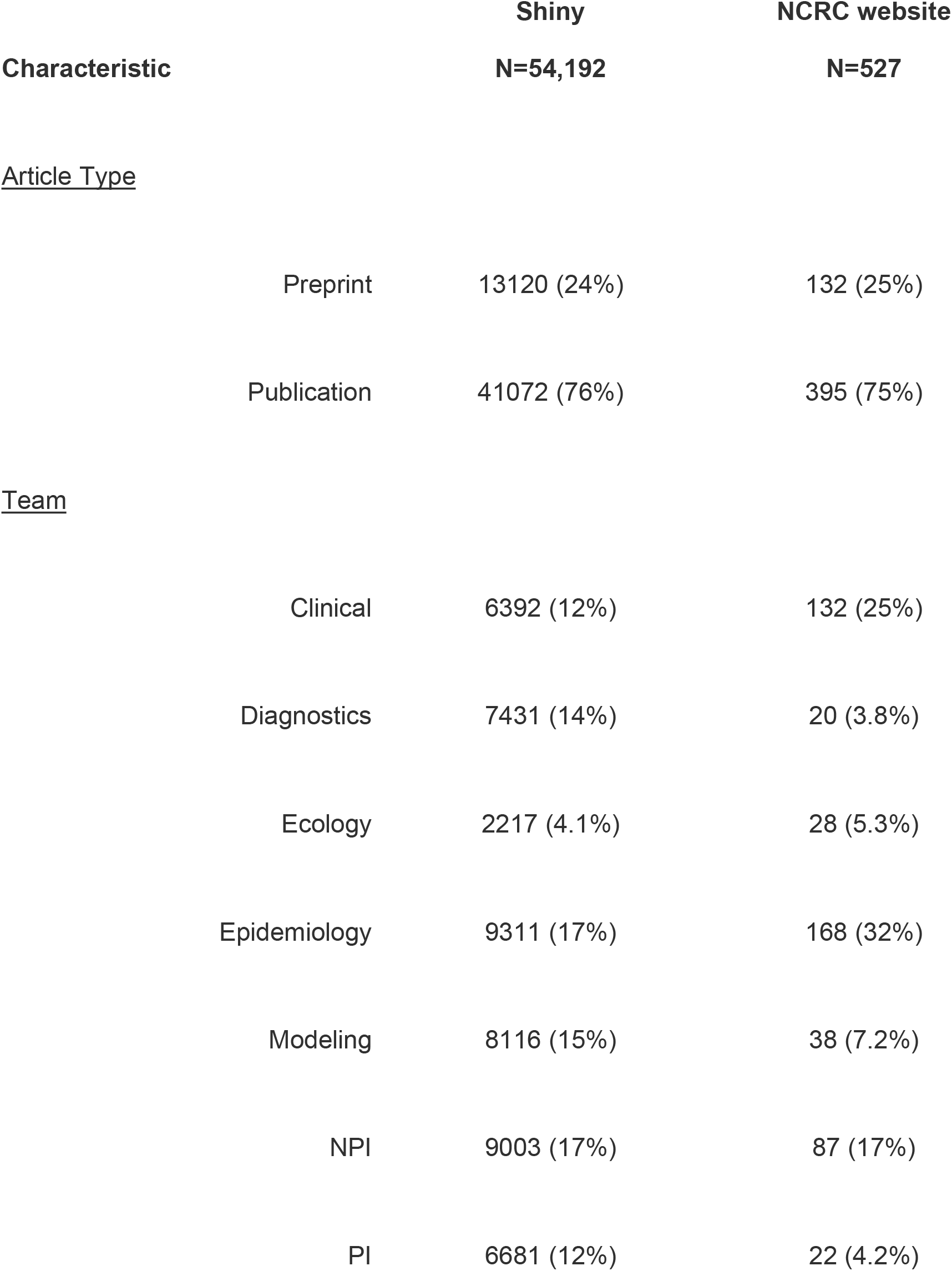
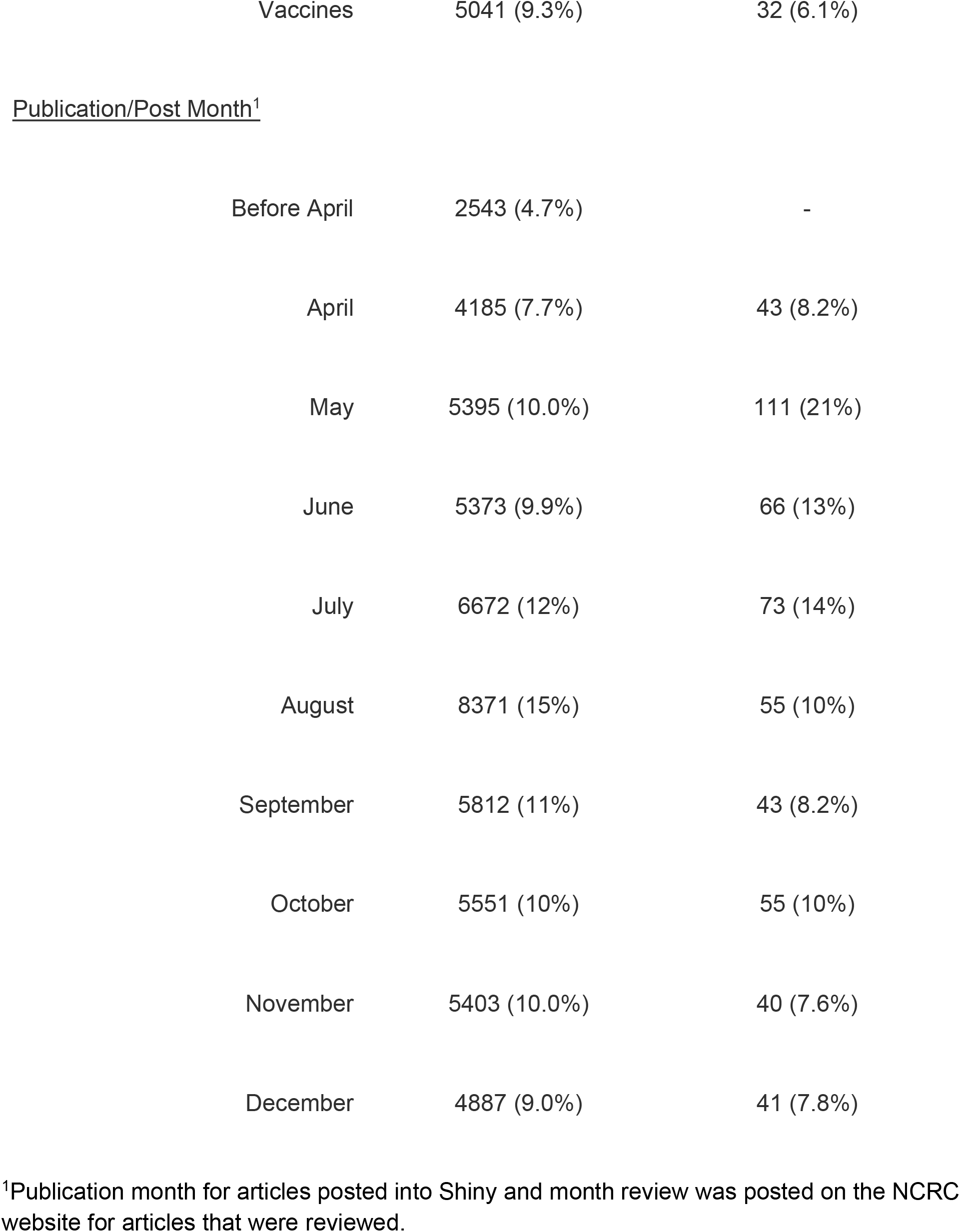
Characteristics of articles pulled into Shiny app (N=51,192) and posted to the Novel Coronavirus Research Compendium (NCRC) website (N=527) from April 6, 2020 to January 1, 2021.

| Characteristic | Shiny<br>N=54,192 | NCRC website<br>N=527 |
| --- | --- | --- |
| <u>Article Type</u> |  |  |
| Preprint | 13120 (24%) | 132 (25%) |
| Publication | 41072 (76%) | 395 (75%) |
| <u>Team</u> |  |  |
| Clinical | 6392 (12%) | 132 (25%) |
| Diagnostics | 7431 (14%) | 20 (3.8%) |
| Ecology | 2217 (4.1%) | 28 (5.3%) |
| Epidemiology | 9311 (17%) | 168 (32%) |
| Modeling | 8116 (15%) | 38 (7.2%) |
| NPI | 9003 (17%) | 87 (17%) |
| PI | 6681 (12%) | 22 (4.2%) |
| Vaccines | 5041 (9.3%) | 32 (6.1%) |
| Publication/Post Month<sup>1</sup> |  |  |
| Before April | 2543 (4.7%) | - |
| April | 4185 (7.7%) | 43 (8.2%) |
| May | 5395 (10.0%) | 111 (21%) |
| June | 5373 (9.9%) | 66 (13%) |
| July | 6672 (12%) | 73 (14%) |
| August | 8371 (15%) | 55 (10%) |
| September | 5812 (11%) | 43 (8.2%) |
| October | 5551 (10%) | 55 (10%) |
| November | 5403 (10.0%) | 40 (7.6%) |
| December | 4887 (9.0%) | 41 (7.8%) |

<sup>1</sup>Publication month for articles posted into Shiny and month review was posted on the NCRC website for articles that were reviewed.

Teams posted 527 total reviews to the NCRC website as of January 1, 2021; 395 (75%) were of peer-reviewed publications and 132 (25%) were of preprints (Table 1, Figure 1). Among 472 articles published after April 1, 2020, the median time from online publication to posting on the NCRC website was 30 days [interquartile range (IQR): 19-46], and this timing was largely consistent by month of review and article status (preprint vs. publication) (Figure 2). Among the 132 preprints reviewed, 73 (55%) were published in a peer-review journal prior to January 27, 2021. The NCRC review of the preprint was posted to the NCRC website a median of 43 days before the publication in a peer-reviewed journal (IQR: 14-93).

**Figure 1.**
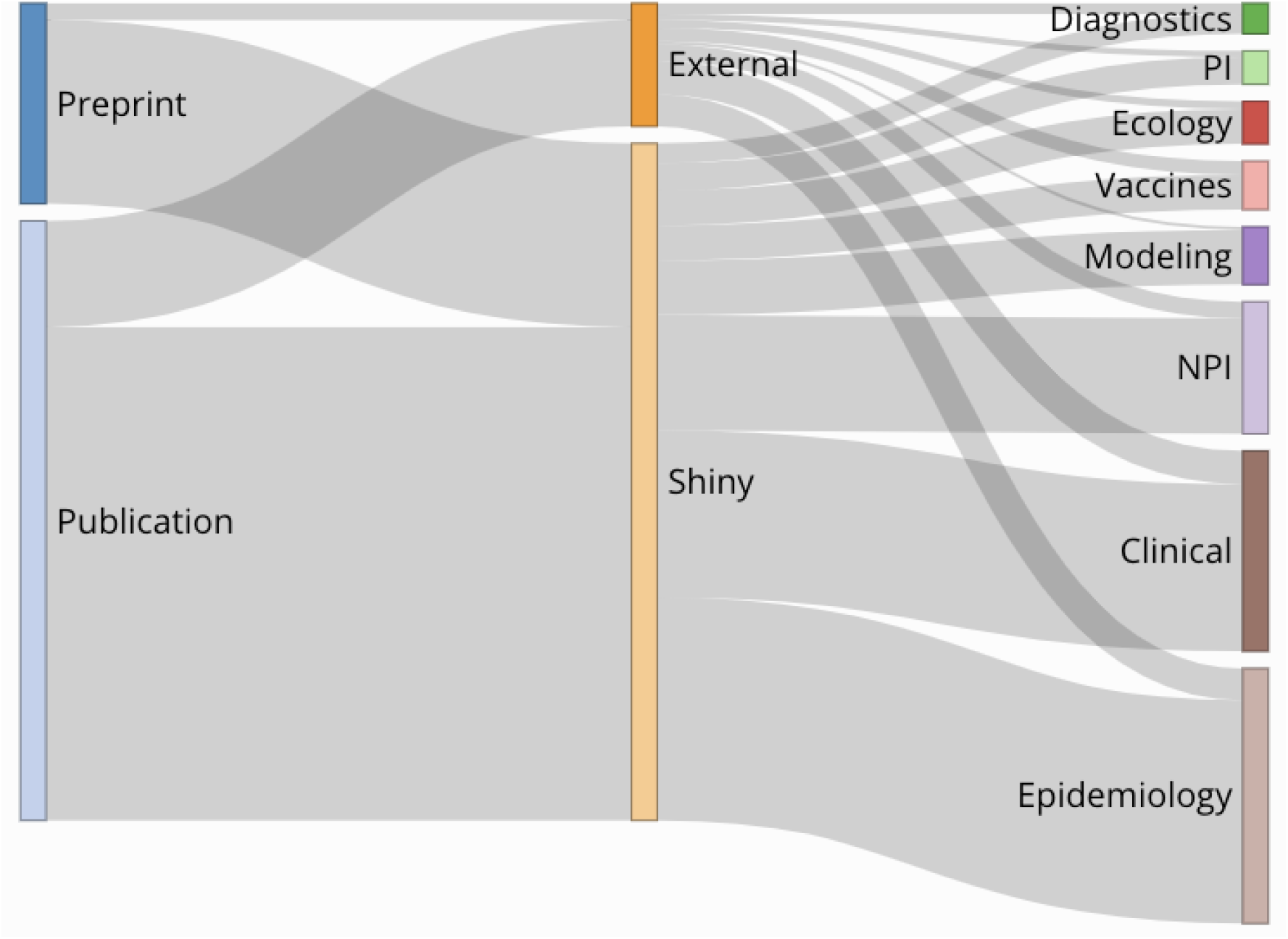
Sankey Diagram of workflow for all reviews published to the Novel Coronavirus Research Compendium (NCRC website p down by source, article type, and primary review group, 1 April 2020 – 1 January 2021 (N=472)

**Figure 2.**
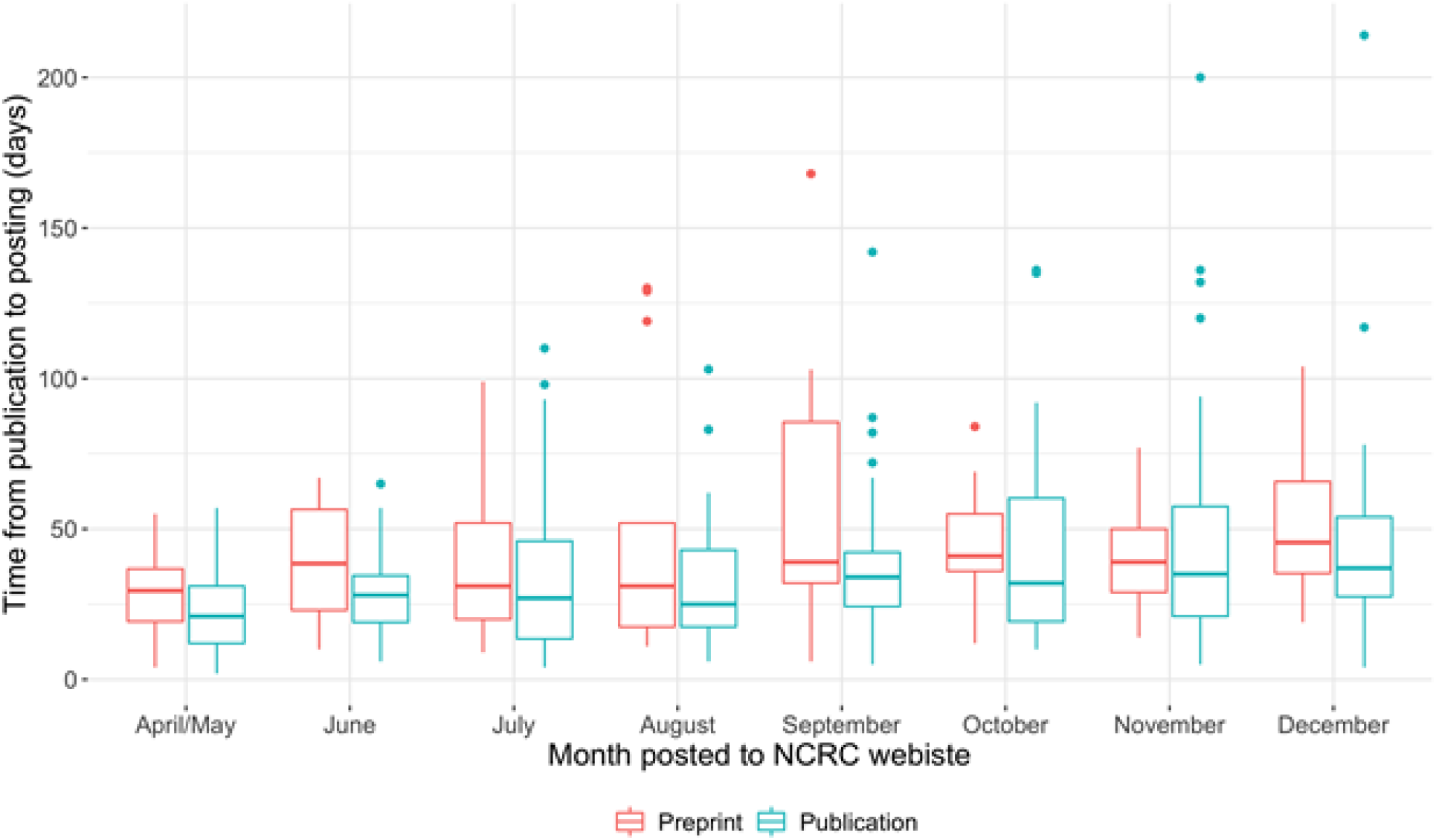
Time gap from posting of preprint or publication of peer-reviewed papers and the date the review was posted on the Novel Coronavirus Research Compendium (NCRC) website, from April 1, 2020 through January 1, 2021 (N=472)

Peer-reviewed publications reviewed by the NCRC (n=395) were published in 102 journals. Over half (62%) of the uploaded reviews of peer-reviewed publications came from seven journals or journal families: Morbidity and Mortality Weekly Report (n=51, 13%), the Lancet family (n=47, 12%), JAMA family (n=36, 9%), Nature family (n=31, 8%), New England Journal of Medicine (n=30, 8%), Clinical Infectious Diseases (n=26, 7%), and Emerging Infectious Diseases (n=22, 6%).

Through January 1, 2021, the NCRC newsletter acquired 1,018 subscribers; the Twitter account had 689,000 impressions; and the website received 112,615 views from 42,174 users, with traffic from 180 countries. The majority of the site’s web traffic was from the United States, United Kingdom, Canada, Spain, Brazil, Germany, and India. Newsletter subscribers include academics, government officials, hospital staff, journalists, nonprofit or for-profit employees, and non-academic research staff.

The NCRC has been covered by media outlets, including *Buzzfeed, Vox*, and *The New York Times*, and scientific magazines such as *Science*. NCRC faculty have also appeared on the “Public Health on Call” podcast produced by the Johns Hopkins Bloomberg School of Public Health to discuss recent reviews.

### Lessons learned

The NCRC represents a large, coordinated, multi-disciplinary effort of technical experts in public health science and clinical medicine dedicated to using their skills to support frontline clinicians and public health practitioners. One of the most important lessons we learned is that our successes required a large team with experience across disciplines. The urgency of the pandemic allowed us to motivate a singular, and often uncompensated, effort to quickly curate and evaluate emerging science. This team-based approach allowed the NCRC to not only code a complex content management system from scratch, but also to cover a wide variety of topics — from transmission across settings, mask-wearing behaviors, and the use of dogs for detecting SARS-CoV-2 to vaccine efficacy against SARS-CoV-2 variants.

In a fast-moving public health crisis, practitioners need the most important and up-to-date evidence quickly to make decisions about clinical care and public health programs. Preprints are an increasingly important pathway for communicating emerging evidence about COVID-19 and SARS-CoV-2. However, there are important risks of using data from preprints to make clinical and public health decisions. As these papers have not yet undergone peer review, they are often more difficult to parse and may contain flaws in their methodology or analyses that could fatally compromise the findings and conclusions. Given the increasing attention from the media on preprints and the urgency to quickly understand the novel coronavirus, the NCRC designed its system to routinely include preprints in the review process. The NCRC’s collaboration with bioRxiv and medRxiv ensures that reviews of preprints are seen by authors and others, like journalists or the public, who might access them directly from the preprint server.

The NCRC’s team of experts aimed to quickly identify popular but flawed articles. Even after peer-review, errors in analysis or overreach in the interpretation of data in research studies may be identified post-publication. The extraordinary speed of publication during the SARS-CoV-2 pandemic may have exacerbated the risk of these errors. Sometimes, errors can give rise to surprising results, and these can garner outsized attention due to the claims they make. On multiple occasions, the NCRC posted reviews to critically evaluate whether the conclusions of a paper were supported by its data and to highlight key methodological shortcomings when applicable to separate actionable evidence from questionable evidence. The NCRC effort to offer critiques in a single place provides practitioners a convenient resource to cut through the noise.

The structure of the NCRC also provides a crucial training opportunity for doctoral students and postdoctoral fellows to understand the connections between academic literature and practice. Students and fellows practice distilling the significance of a paper and translating science into accessible language for decision makers. In addition to providing mentorship on science communication, faculty also meet regularly with students and fellows to reflect on which papers should be included on the NCRC website. The curation process helps to hone the skills of students and fellows to understand the current landscape, gaps in knowledge, and which research questions are indispensable for moving the evidence base forward.

Despite these strengths, the sustainability of such a large effort is unclear. In particular, the NCRC has been unable to identify long-term funding to cover the costs of faculty time dedicated to the project and this poses a risk to the future of the endeavor. The time lag between paper publication and our reviews has been a concern, since it has reduced our ability to contribute to discussions about evidence in real time. These delays had multiple contributors, including the lags between publication and indexing in PubMed, but a lack of salary support for the effort also contributed. Our ability to measure the impact of the NCRC on changes to knowledge or practice among our target audience has been limited, for multiple reasons. Any impact measurement would almost certainly require interviews with members of our target audience, who currently have no time to spare from pandemic response to participate in such endeavors. Additionally, the NCRC team has been stretched to keep up with the literature leaving little time to focus on impact assessments, though future work focused on outcome assessment should be considered. Nevertheless, continued utilization of the reviews, reflected in growing newsletter subscribers and increases in webpage views, and informal conversations with colleagues in our target audience suggest that the NCRC is a valued resource.

As the COVID-19 pandemic has emphasized the necessity of collaboration between public health researchers and practitioners to save lives, we believe that the NCRC is one model of how this collaboration can be successful. Since its inception, we have conceptualized the NCRC as a critical training opportunity for the next generation of public health and clinical scientists in knowledge translation to improve decision making. The NCRC approach could be useful to improve timely translation of data to action for other public health problems where lags in knowledge translation persist, particularly if funding were available to support the effort. By using this model, we have improved the ability of public health officials and clinicians to respond to the COVID-19 pandemic and built systems and capacity that can be applied to solving other public health problems, including the next pandemic.

## Data Availability

All data are either publicly available or available upon request.

## Footnotes

1 Based on results from a search on all papers published on these topics since 1940.

